# Altered correlation of concurrently recorded EEG-fMRI connectomes in temporal lobe epilepsy

**DOI:** 10.1101/2022.09.01.22279214

**Authors:** Jonathan Wirsich, Giannina Rita Iannotti, Ben Ridley, Elhum A Shamshiri, Laurent Sheybani, Frédéric Grouiller, Fabrice Bartolomei, Margitta Seeck, François Lazeyras, Jean-Philippe Ranjeva, Maxime Guye, Serge Vulliemoz

## Abstract

Whole brain, large-scale functional connectivity networks or connectomes have been characterized on different temporal and spatial scales in humans using EEG and fMRI. Whole brain epileptic networks have been investigated with both EEG and fMRI, but due to the different acquisition approaches it is unclear to what extent those results can be related. In consequence clinical research in epilepsy would profit from a unified multimodal functional connectome description as a linking framework to better map underlying brain function and pathological functional networks. In this study we aim to characterize the spatial correlation between EEG and fMRI connectivity in temporal lobe epilepsy.

From two independent centers, we acquired resting-state concurrent EEG-fMRI from a total of 35 healthy controls and 34 TLE patients (18 right TLE and 16 left TLE). Data was projected into the Desikan brain atlas (mean BOLD activity for fMRI and source reconstruction for EEG). Whole brain functional connectivity from fMRI (Pearson correlation) and EEG (corrected imaginary part of the coherency) were correlated for all subjects.

In healthy controls, average EEG and fMRI whole-brain connectivity was moderately correlated (r∼0.3). For both imaging centers, correlation between EEG and fMRI whole brain connectivity was increased in rTLE when compared to controls for lower frequency bands (EEG-delta, theta and alpha). Conversely correlation between EEG and fMRI connectivity of lTLE patients was decreased in respect to healthy subjects (EEG-beta vs. fMRI connectivity only). While the alteration of the EEG-fMRI correlation in rTLE patients could not be related to a local effect, in lTLE patients it was locally linked to the Default Mode Network.

We demonstrated, using two independent datasets, that EEG and fMRI connectivity is correlated for both healthy subjects and patients. The increased correlation of EEG and fMRI connectivity in rTLE patients vs. controls and decreased correlation in lTLE patients vs. controls suggests a differential organization of mono-lateral focal epilepsy of the same type, which needs to be considered when comparing fMRI to EEG connectivity. It also demonstrates that each modality provides distinct information, highlighting the benefit of multimodal assessment in epilepsy. The observed property of distinct topological patterns depending on the lateralization of the epilepsy could be taken into account when clinically defining the epileptic focus of patients.

## Introduction

It now is consensus that multimodal integration of whole-brain imaging facilitates the clinical exploration of brain pathology. However, it is yet an open question how multimodal measures of pathological brain networks can help in epilepsy to guide clinical diagnosis, treatment and brain surgery (Zijlmans et al., 2019). While epileptic phenomena are clinically characterized by altered brain rhythms and paroxysmal local discharges, recorded using the electroencephalogram (EEG), more widespread whole brain functional alteration linked to epilepsy has been characterized by functional MRI (fMRI) (Centeno and Carmichael, 2014). In a clinical context the fast dynamics of EEG and the finer spatial resolution of fMRI can be used to investigate the hemodynamic changes correlated with epileptic spikes in order to obtain an improved spatial characterization of the epileptogenic network (Vulliemoz et al., 2009).

To investigate whole brain functional network alterations associated with epilepsy, fMRI (Bettus et al., 2009; Ridley et al., 2015) and EEG (Coito et al., 2015) have been successfully applied, but it remains unclear how results extracted from different modalities can be used together in a meta-analysis (Slinger et al., 2022; van Diessen et al., 2014). To translate basic research results derived from complex fMRI connectivity graph models into clinical management of patients with epilepsy, it is indispensable to better understand the correspondence between EEG and fMRI connectivity.

In healthy subjects, moderate correlations between EEG and fMRI functional connectivity (FC_fMRI_ and FC_EEG_) exist (Deligianni et al., 2014; Wirsich et al., 2021) and EEG and fMRI connectivity dynamics are linked to each other (Wirsich et al., 2020b) while parts of the FC_EEG_ and FC_fMRI_ provide complimentary information (Wirsich et al., 2020a, 2017). Being able to extract both commonalties and discrepancies between FC_fMRI_ and FC_EEG_ is encouraging as they point in the direction that whole-brain networks extracted from clinical EEG can be generally used instead of a more expensive assessment with fMRI. As such mapping FC_EEG_ and FC_fMRI_ into one graph space provides a framework to translate fMRI findings into the clinical setting of EEG recordings. For this purpose, it is necessary to understand if the relationship between EEG and fMRI is altered when comparing healthy subjects and patients with epilepsy. Alterations between healthy and pathological networks in electrophysiology and hemodynamics are complex and specific alterations of the EEG-fMRI relationship have been reported in combination with several EEG-frequency bands while the reproducibility of those individual studies remains unclear (Centeno and Carmichael, 2014). An unaltered FC_EEG_ the FC_fMRI_ relationship would suggest that recording a single modality may be enough to characterize functional connectivity alterations in epilepsy, while a changed relationship would highlight the importance of multimodal exploration (Forsyth et al., 2019).

In this study we sought to characterize the spatial correlation between whole-brain FC_EEG_ and FC_fMRI_ in order to understand if the crossmodal mapping of FC_EEG_ and FC_fMRI_ is modified in patients with epilepsy as compared to healthy controls. This will close the knowledge gap of how FC_fMRI_ and FC_EEG_ studies compare in focal epilepsies. The advantage of this approach is that the exact topology of reorganization is irrelevant: the spatial correlation of whole brain EEG and fMRI connectivity will measure the topological alteration of networks that generalize across the patient group while omitting local patient-specific functional reorganization. We aimed to assess the reproducibility of our results by using two independently recorded EEG-fMRI datasets.

## Methods

### Participants and EEG-fMRI data acquisition

We included patients with drug resistant focal temporal lobe epilepsy with clear unilateral epileptic focus (clinically defined by combined information from imaging, interictal epileptiform discharges (IEDs) and seizure onset) alongside with healthy controls. To do so, we retrospectively used data from two independent centers using a 256-channel EEG setup in a 3T scanner (dataset will be referenced as 256Ch-3T) and a 64-channel EEG setup in a 3T scanner (dataset 64Ch-3T). We included data of resting-state concurrent EEG-fMRI acquisitions in a total of 35 healthy controls (64-3T: 14 and 256-3T: 21) and a total 34 patients diagnosed with drug-resistant epilepsy of the temporal lobe (TLE, 64-3T: n=11 and 256-3T n=23 / distribution of left and right TLE: rTLE n=18 and lTLE n=16).

#### 256Ch-3T

21 healthy subjects (7 females, mean age: 32, age range 24–47) with no history of neurological or psychiatric illness and 23 TLE patients (14 females, mean age: 34, age range 18-60, 13 lTLE and 10 rTLE) were recorded. Ethical approval was given by the local Research Ethics Committee (Commission Cantonale d’Ethique, Genève) and informed consent was obtained from all subjects. The control group has been previously analyzed in (Wirsich et al., 2021).

For patient and control group a variable time period of resting-state simultaneous EEG-fMRI data were acquired. In order to have a consistent recording length within the dataset we only analyzed the first 4min58s of each dataset (see SI Table 1). Subjects were asked not to move, to remain awake and keep their eyes closed during the resting-state scan. MRI was acquired using a 3 Tesla MR-scanner (Siemens Magnetom Trio / Siemens Magnetom Prisma, update of clinical scanner during protocol see SI Table 1). The fMRI scan comprised the following parameters: GRE-EPI sequence, TR=1980/1990/2000 ms (for details see update of clinical scanner during protocol see SI Table 1), TE=30 ms, 32 slices, voxel size 3 × 3 × 3.75mm^3^, flip angle 90°. Additionally, an anatomical T1-weighted image was acquired (176 sagittal slices, 1.0 × 1.0 × 1.0 mm, TA=7 min). EEG was acquired using a 258-channel MR-compatible amplifier (Electrical Geodesic Inc., Eugene, OR, USA, sampling rate 1 kHz), including 256 electrodes (Geodesic Sensor Net 256, referenced to Cz) and 2 ECG electrodes (bipolar montage, placed on the chest, crossing the heart). The scanner clock was time-locked with the amplifier clock (Mandelkow et al., 2006). An elastic bandage was pulled over the subjects’ head and EEG cap to assure the contact of electrodes on the scalp. The MR-compatible amplifier was positioned to the left of the subject and EEG and ECG cables were passed through the front end of the bore.

#### 64Ch-3T

14 healthy subjects (5 females, mean age: 31, age range 20-55) with no history of neurological or psychiatric illness and 11 TLE patients (6 females, mean age: 37, age range 22-54, 3 lTLE and 9 rTLE) were recorded. Ethical approval was given by local Research Ethics Committee (Comité de Protection des Personnes (CPP) Marseille 2) and informed consent was obtained from all subjects. Data of the control group has been previously analyzed in Wirsich et al. (2020a, 2017).

In each subject one run of 21min resting-state simultaneous EEG-fMRI was acquired. Subjects were asked not to move and to remain awake and keep their eyes closed during the resting-sate scan. MRI was acquired using a 3 Tesla MR-scanner (Siemens Magnetom Verio 3T). The fMRI scan comprised the following parameters: GRE-EPI sequence, TR=3600 ms, TE=27 ms, 50 slices, voxel size 2 × 2 × 2.5mm, flip angle 90°, total of 350 vols. Additionally, an anatomical T1-weighted image was acquired (208 sagittal slices, 1.0 × 1.0 × 1.0mm, TA=6min27s).

EEG was acquired using a 64-channel MR-compatible amplifier (BrainAMP MR – Brain Products, Munich, Germany, sampling rate 5 kHz), 64 electrodes (referenced to FCz, 1 ECG electrode placed on the chest above the heart). The scanner clock was time-locked with the amplifier clock (Mandelkow et al., 2006). The amplifier was placed as far as possible behind the scanner and the connector cables were fixed with sandbags to avoid distortions due to mechanical vibrations of the scanner.

## Data Processing

Data preprocessing was carried out as described in Wirsich et al. (2021).

### Brain Parcellation

We used the Freesurfer toolbox (Fischl, 2012) to process the T1-weighted images (recon-all, v6.0.0 http://surfer.nmr.mgh.harvard.edu/) by performing non-uniformity and intensity correction, skull stripping and gray/white matter segmentation. The cortex was parcellated into 68 cortical regions according to the Desikan(-Killiany) atlas (Desikan et al., 2006).

### fMRI Processing

Slice timing correction was applied to the fMRI timeseries. This was followed by spatial realignment both using the SPM12 toolbox (revision 7475; http://www.fil.ion.ucl.ac.uk/spm/software/spm12). The T1 images of each subject and the Desikan atlas were coregistered to the fMRI images (FSL-FLIRT 6.0.2, https://fsl.fmrib.ox.ac.uk/fsl/fslwiki, (Jenkinson et al., 2012)). We extracted signals of no interest such as the average signals of cerebrospinal fluid (CSF) and white matter from manually defined regions of interest (ROI, 5 mm sphere, Marsbar Toolbox 0.44, http://marsbar.sourceforge.net) and regressed them out of the BOLD timeseries along with 6 rotation, translation motion parameters and global gray matter signal (Wirsich et al., 2017). Then we bandpass-filtered the timeseries at 0.009–0.08 Hz (Power et al., 2014). Like in Wirsich et al. (2021), we scrubbed the data using frame wise displacement (threshold 0.5 mm, by excluding the super-threshold timeframes) as defined by Power et al. (2012).

### fMRI connectivity measures

Average timeseries of each region was then used to calculate FC_fMRI_ by taking the pairwise Pearson correlation of each regions’ cleaned timecourse (see schema Fig 1). The final connectivity matrix was constructed from the unthresholded values of the Pearson correlation.

**Fig 1:**
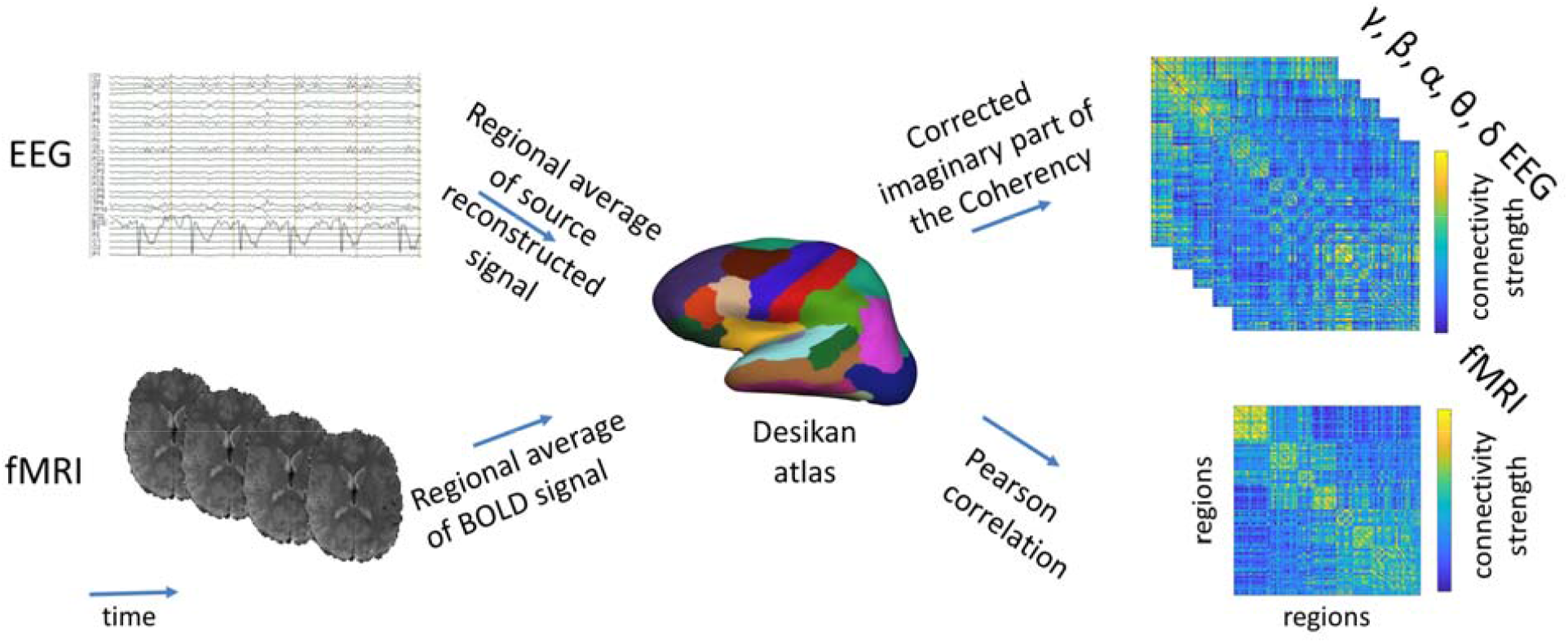
Overview on the construction of EEG and fMRI connectomes. EEG and fMRI data were parcellated into the 68 regions of the Desikan atlas (coregistered to each subject’s individual T1) as follows: For fMRI, the BOLD signal timecourse was averaged over the voxels in each region for each subject. The Pearson correlation of the region averaged fMRI-BOLD timecourse was calculated to build a function connectivity matrix/connectome (FC_fMRI_). For the EEG, the signal of each sensor was source reconstructed to the cortical surface (15,000 solution points) using the Tikhonov-regularized minimum norm. Then, the timecourses of the solution points were averaged per cortical region. The corrected imaginary part of the coherency (ciCoh) of averaged EEG source signals were used to calculate FC_EEG_ for each subject (Figure adapted from (Wirsich et al. 2021). Please refer to the methods for a detailed description of each step).

### EEG Processing

EEG data was preprocessed individually for the different setups:

#### 256Ch-3T

EEG was corrected for the scanner gradient artifact using template subtraction with optimal basis set and adaptive noise cancelation (Allen et al., 2000; Niazy et al., 2005), followed by pulse-related artifact template subtraction (Allen et al., 1998) using in-house code Matlab code for ballistocardiogram peak detection as described in Iannotti et al. (2015). Electrodes placed on the cheeks and in the face were excluded from data analysis resulting in a final set of 204 used electrodes. This was followed by manual ICA-based denoising (for manual removal of gradient and pulse artifact residuals, eye-blinks, muscle artifacts, infoMax, runICA-function EEGLab revision 1.29 (Bell and Sejnowski, 1995; Delorme and Makeig, 2004)).

#### 64Ch-3T

The Brain Vision Analyzer 2 software (Brain Products, Gilching, Germany) was used for the following processing steps. EEG was corrected for the scanner gradient artifact using template subtraction, adaptive noise cancelation and downsampling to 250 Hz (Allen et al., 2000) followed by pulse-related artifact template subtraction (Allen et al., 1998). Then ICA-based denoising (for manual removal of gradient and pulse artifact residuals, eye-blinks and muscle artifacts, Fast ICA restricted mode with probabilistic sphering) was carried out. Data was segmented according to one TR of the fMRI acquisition (TR=3600ms). The segments with obvious movement artifacts were semi-automatically excluded from further analysis (Wirsich et al., 2017). Finally the data was bandpass-filtered the signal at 0.3-70 Hz.

#### Both datasets

A trained neurologist (L.S.) visually inspected all EEG data to mark interictal epileptiform discharges (IEDs), IED segments were not removed but were used as a covariable in our analysis. Cleaned EEG data was imported and analyzed with Brainstorm software (Tadel et al., 2011), which is documented and freely available under the GNU general public license (http://neuroimage.usc.edu/brainstorm, version 15th January 2019).

#### 256Ch-3T

(the following steps where already carried out in the Brain Vision Analyzer software for 256Ch-3T data): Data was bandpass-filtered at 0.3–70 Hz. Data was segmented according to one TR of the fMRI acquisition (TR=1980-2000ms, see SI Table 1). In order to minimize effect of head motion EEG epochs containing motion were automatically detected if the signal in any channel exceeded the mean channel timecourse by 4 standard deviations. Then the whole timecourse was also visually inspected to exclude all motion segments from further analysis (Wirsich et al., 2021).

#### Both datasets

Channels that remained artefactual were removed from the analysis (without interpolation). Electrode positions and T1 were coregistered by manually aligning the electrode positions onto the electrode artifacts visible in the T1 image. A forward model of the skull was calculated based on the individual T1 image of each subject using the OpenMEEG BEM model, (Gramfort et al., 2010; Kybic et al., 2005). The EEG signal was re-referenced to the global average and projected into source space (15,000 solution points on the cortical surface) using the Tikhonov-regularized minimum norm (Baillet et al., 2001) with the Tikhonov parameter set to 10% (Brainstorm 2018 implementation, with default parameters: assumed SNR ratio 3.0, using current density maps, constrained sources normal to cortex with signs flipped into one direction, depth weighting 0.5/max amount 10). Finally, the source activity of each solution point was averaged in each cortical region of the Desikan atlas.

### EEG connectivity measures

For each epoch (each TR) the corrected imaginary part of the coherency (ciCoh, (Ewald et al., 2012; Nolte et al., 2004)) of the source activity was calculated between each region pair (cortical regions only: Desikan atlas - 68 regions) using bins of 2 Hz frequency resolution (Wirsich et al., 2021) (Brainstorm implementation, version 15–01–2019; imaginary part was corrected by the real part of the coherence 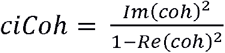 (Ewald et al., 2012), significance of each connectivity value was determined according to Schelter et al. (2006), connections with p>0.05 were set to 0). The 2 Hz bins were averaged for 5 canonical frequency bands: delta (δ 0.3–4 Hz), theta (θ 4–8 Hz), alpha (α 8–12 Hz), beta (β 12–30 Hz), and gamma (γ 30–60 Hz). The segments were then averaged across time for each subject to one FC_EEG_ matrix (see schema Fig 1).

## Connectivity Analysis

### Split-half and cross-dataset spatial correlation

Spatial similarity of monomodal FC was assessed by correlating the split-half averages of the upper triangular of the connectivity matrix of each dataset and group. To do so, each group was randomly split into two equally sized datasets and the correlation between the two split-averaged matrices was calculated for multiple iterations. We report the correlation averaged over each split-half iteration (5000 iterations or in the case of group sizes n<16 we calculated all possible combinations to split the dataset into two parts). As those split-half correlations depend on the group size, the results should be only used to qualitatively assess the data and compare them to the results of Wirsich et al. (2021), but not to assess differences between controls and patients. Monomodal cross-dataset spatial correlation was assessed by correlating group averages of each dataset with the respective participant group in the other dataset.

### Network based statistics of monomodal measures

With the goal to better understand if the EEG and fMRI connectomes are altered across groups due to local and monomodal shifts of connectivity we used network-based statistics (Zalesky et al., 2010) on each modality. For FC_EEG_ this was done for each frequency band. In detail we built 6 linear models with FC_fMRI_ (Fisher z-transformed), FC_EEG-δ_, FC_EEG-θ_, FC_EEG-α_, FC_EEG-β_ and FC_EEG-γ_ as response variables, group label as regressor of interest and age, sex and dataset site as regressor of non-interest. We tested for monomodal network changes between controls and patients by applying the following contrasts: controls>rTLE, controls>lTLE, controls<rTLE and controls<lTLE (one-sided t-test, connection level threshold T=2, NBS-corrected threshold adapted to 6 models p<0.05/6∼0.0083).

### Crossmodal spatial EEG-fMRI connectivity correlation

Crossmodal spatial correlations between FC_EEG_ and FC_fMRI_ of each group-averaged connectivity matrices were calculated. To test if the crossmodal correlation of rTLE and lTLE patients was different to the one of healthy controls, we built a distribution of 5000 averaged matrices by randomly switching the group labels (Wirsich et al., 2016). Previously, we demonstrated in healthy controls that the spatial relationship of EEG-fMRI connectivity can be robustly extracted when averaging around 7-12 subjects (Wirsich et al., 2021). This excellent reproducibility of averaged resting state recordings was also recently demonstrated on large fMRI datasets (n>1000, r>0.9 for average connectomes with n>10, see supplementary Figure 17 within (Marek et al., 2022)). The number of lTLE patients in the 64Ch3T-dataset was only n=3, and in consequence we did not carry out any group-averaged analysis using only subjects restricted this group/dataset combination.

To understand how the crossmodal correlation is influenced by age, sex, epilepsy duration (as epilepsy onset and duration are correlated, we decided to use only duration), etiology and IEDs we generated several bootstrapped distributions (with replacement, Matlab bootstrap function, 1000 iterations) of the average EEG-fMRI correlation. This bootstrapping method will generate an average value for each EEG and fMRI connection that can be used to generate a bootstrapped FC_EEG_-FC_fMRI_ correlation alongside with subject specific variables such as age (e.g., one bootstrap iteration might result in an EEG-fMRI correlation of r=0.3 an average female/male ratio of 0.4 and an average age of 33.1 while the next iteration will end up with r=0.35, ratio=0.45 and average age of 34.2). Each iteration of the bootstrapped averages were then used in three linear models to identify the relationship of each bootstrapped, averaged variable to the bootstrapped, averaged EEG-fMRI correlation (Model I: controls-lTLE patients: r(EEG-fMRI) ∼ age + sex + group-label + dataset-site, Model II: controls-rTLE patients: r(EEG-fMRI) ∼ age + sex + group-label + dataset-site, Model III: patients: r(EEG-fMRI) ∼ age + sex + epilepsy-duration + isHS + recorded IEDs per minute + group-label + dataset-site; isHS=binary dummy variable coding for hippocampal sclerosis or not, for etiology distribution other than HS see patient description in **Error! Reference source not found**., epilepsy-duration is coded in full years, dataset-site=dummy variable coding for 256Ch-3T or 64Ch-3T dataset). To test for significance of the contribution to the EEG-fMRI connectivity correlation, the T-value of each coefficient/variable in the linear model was compared to a null-model that bootstrapped (1000 iterations with replacement) the averages of the same model having the target variable permuted across the dataset (e.g. group-labels switched between lTLE and controls, 5000 iterations).

### Spatial subnetwork contribution to the EEG-fMRI connectivity correlation

To better understand the spatial contributions to cross modal correlations of the whole brain we split the FC-matrices into subnetworks of the 7 ICNs (Visual, Somato-motor, Ventral Attention, Dorsal Attention, Front-Parietal, Limbic and Default Mode) as defined by Yeo et al. (2011). For each subdivision we individually assessed the crossmodal correlation of the intra-subnetwork connections in order to statistically compare the difference between controls and patients (lTLE≠controls / rTLE≠controls, permutation test of group labels, 5000 iterations). Equally we assessed the contribution of each connection to the total crossmodal correlation (Colclough et al., 2016; Wirsich et al., 2021). In brief the relative spatial contribution c of each connection i is given by: 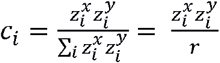 with 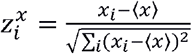 and 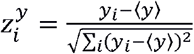 given the Pearson correlation coefficient of two vectors x and y: 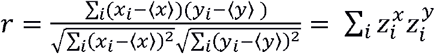. This spatial contribution was statistically compared between patients and controls (direction of the test was chosen according to the results of the FC_EEG_-FC_fMRI_ correlation lTLE<controls / rTLE>controls, permutation of group labels, 5000 iterations).

All p-values are reported uncorrected, and the corresponding Bonferroni-correction threshold is explicitly stated alongside each individual analysis.

## Results

### Behavioral

No significant difference in head movement measured by framewise displacement (Power et al., 2012) was observed between rTLE vs. controls and lTLE vs. controls, (two-sided ttest, all p>0.05 uncorrected).

### Monomodal split-half and cross-dataset correlation

Intra-group monomodal consistency of the subject groups (split by dataset-site and control, lTLE and rTLE patient group) was accessed by randomly splitting the dataset into two equally sized parts (5000 iterations or all combinations in case the number of subjects in the group was n<16) and spatially correlating the FC_EEG_ and FC_fMRI_ matrices. The split-half correlation of FC_fMRI_ ranged from r=0.88 (controls, dataset 64Ch3T) to r=0.62 (rTLE patients, dataset 256Ch3T). The FC_EEG_ split-half correlation ranged from r=0.82 (FC_EEG-β_, controls, dataset 256Ch3T) to r=0.28 (FC_EEG-γ_, rTLE patients, dataset 64Ch3T, for all results see SI Table 2).

### Monomodal contributions

When comparing the monomodal connectivity we could not find any significant differences between rTLE and controls and lTLE and controls for FC_fMRI_ and FC_EEG_ (controls>rTLE, controls>lTLE, controls<rTLE and controls<lTLE, one-sided t-test, connection level threshold T=2, NBS-corrected threshold adapted to 6 models p<0.05/6∼0.0083)

### EEG-fMRI correlation

In line with Wirsich et al. (2021, 2017) healthy controls moderately (r∼0.3-0.4) correlated in the 256Ch-3T and 64Ch-3T dataset. EEG-fMRI correlation was also moderately correlated (r∼0.3-0.4) in both patient groups (Fig 2, SI Table 3).

**Fig 2:**
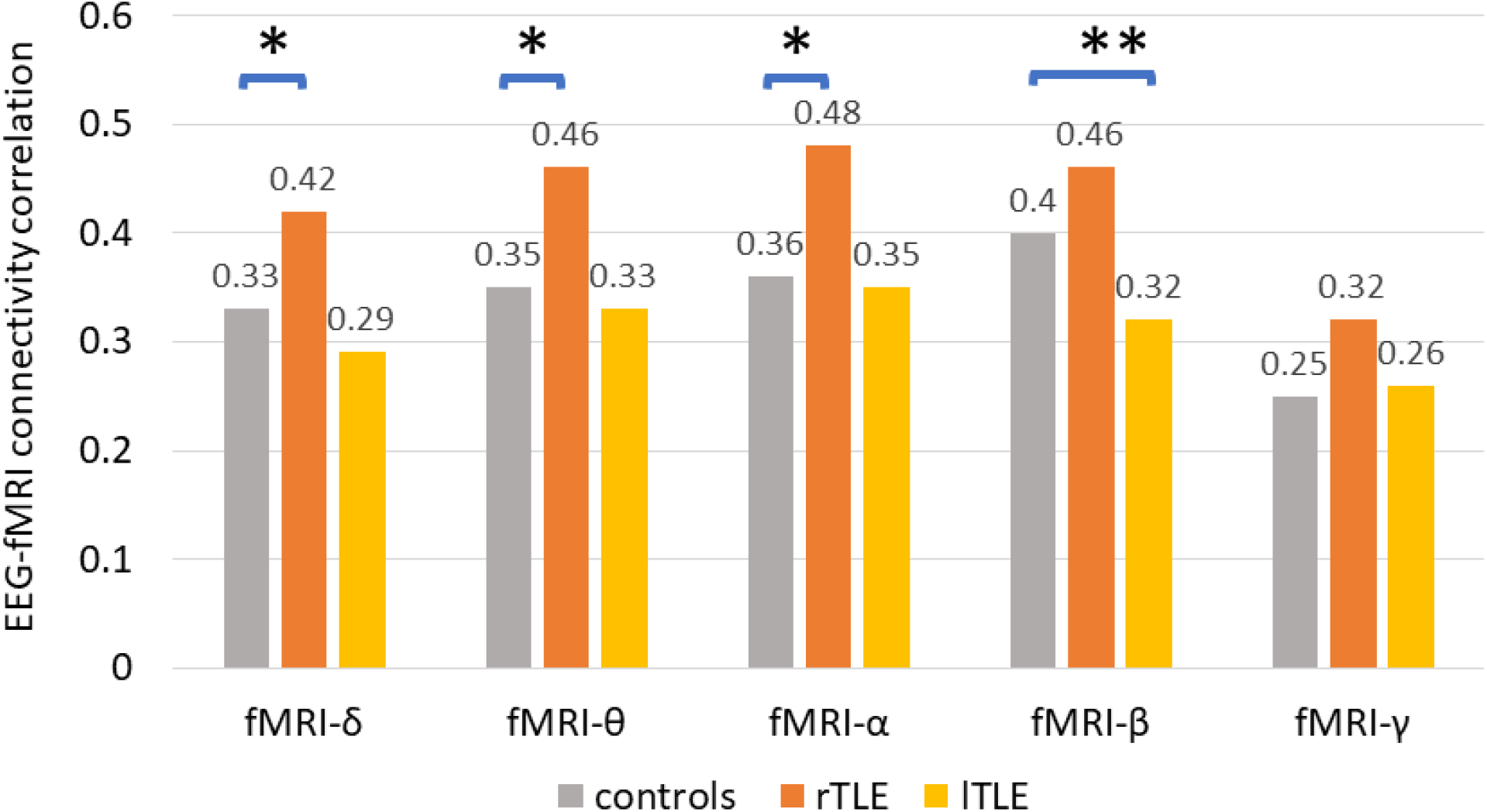
Crossmodal correlation between group-averaged FC_EEG_ and FC_fMRI_ (pooled across centers according to (Wirsich et al., 2021)) using the Desikan atlas (*rTLE patients > controls ** lTLE patients < controls: Bonferroni threshold: p<0.05/5=0.01, permutation test with 5000 iterations, for all results see SI Table 3)

As compared to healthy controls, crossmodal correlation of rTLE patients was increased in FC_EEG-δ_, FC_EEG-θ_ and FC_EEG-α_ (corrected Bonferroni threshold: p<0.05/5=0.01, see Fig 2 and Fig 3A). For lTLE patients we observed significantly decreased beta as compared to healthy controls (r(FC_fMRI_,FC_EEG-β_): lTLE< controls, corrected Bonferroni threshold: p<0.05/5=0.01, see Fig 2 and Fig 3A).

**Fig 3:**
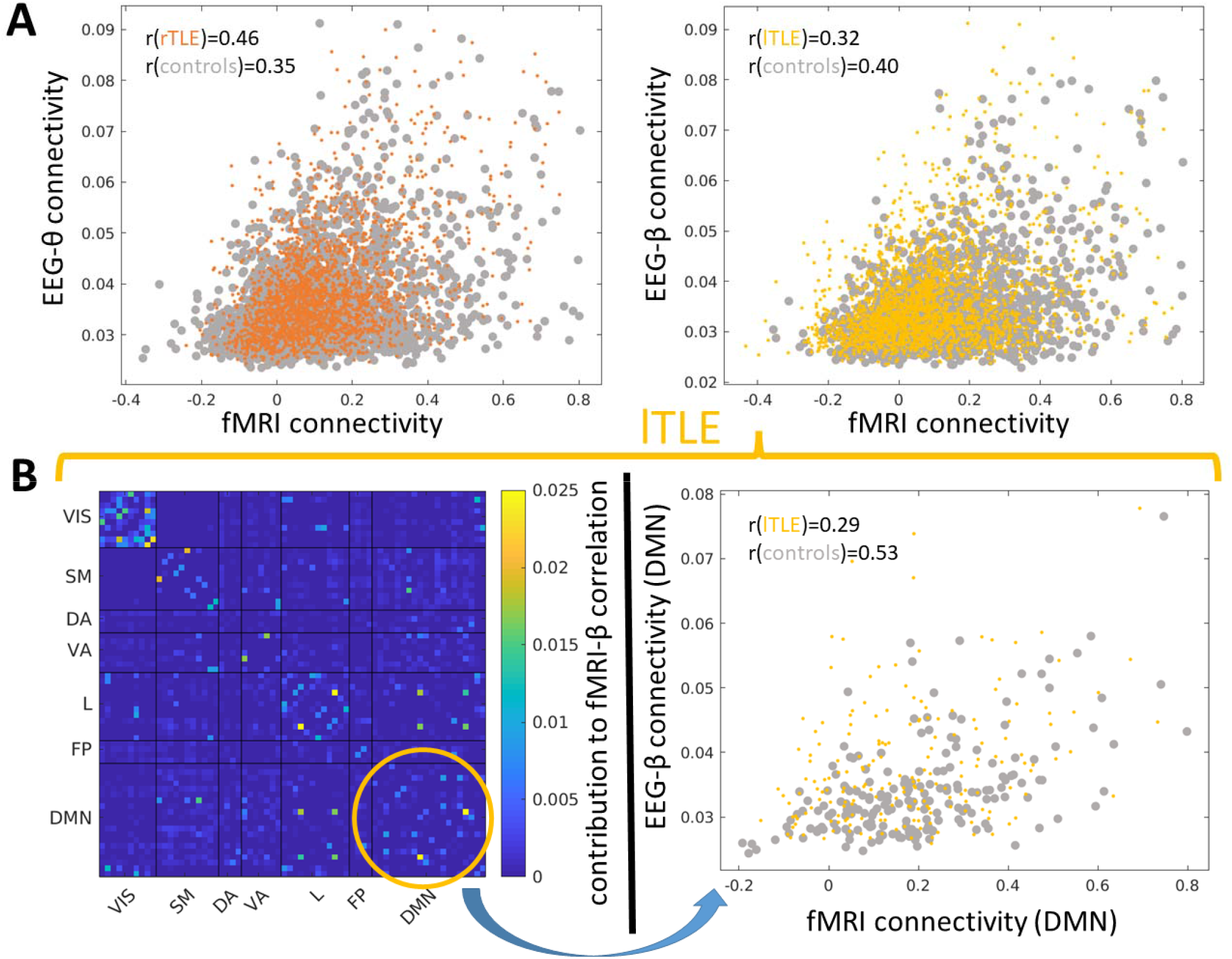
Scatter plots of all pairwise FC_fMRI_ and FC_EEG_ connection strengths (each point samples the FC_EEG_ and FC_fMRI_ connectio strength of one region pair of the group averaged FC). A (left side): Significant FC_EEG_-FC_fMRI_ correlation differences in controls and rTLE patients in the θ-band and A (right side): Controls and lTLE patients in the β-band. B (left side): Spatial contribution to FC_fMRI_-FC_EEG_ correlation of lTLE patients (yellow circle depicts DMN network that was significantly decreased in lTLE patients as compared to healthy controls). B (right side) Scatter plot of FC_fMRI_ and FC_EEG_ connection strengths in the DMN which are significantly less correlated in lTLE patients as compared to healthy controls; we did not find any significant local alterations of the crossmodal relationship when comparing rTLE patients to healthy controls (see SI Table 3); VIS: Visual, SM: Somato-Motor, DA: Dorsal Attention, VA: Ventral Attention, L: Limbic, FP: Fronto-Parietal, DMN: Default Mode Network

When combining all subjects to a grand average (patient and control group) the multimodal correlation peaks at r∼0.40 (except γ: r=0.33, see 1^st^ row of SI Table 3) as observed in Wirsich et al. (2021). As we have previously demonstrated, adding more healthy controls will generally increases the correlation (Wirsich et al., 2021) the higher correlation of rTLE (n=17) compared to all pooled subjects (n=35) makes the possibility that the result is driven by a random higher SNR of rTLE patients very unlikely (see SI Table 3).

### Bootstrapping the group averaged FC_fMRI_-FC_EEG_ correlation

Using bootstrapped group averages in a linear model in order to analyze how different resampling-iterations with replacements change the average EEG-fMRI correlation we observed that:

1. in a model including controls and rTLE patients (controlling for age, sex and dataset-site): FC_fMRI_-FC_EEG-δ_/-FC_EEG-θ_/-FC_EEG-α_/-FC_EEG-β_ correlation was significantly increased for rTLE patients as compared to healthy controls (p<0.05/5=0.01, Bonferroni corrected).
2. in a model including controls and lTLE patients (controlling for age, sex and dataset-site): FC_fMRI_-FC_EEG-β_ correlation was significantly decreased (uncorrected only) for rTLE patients as compared to healthy controls (uncorrected p<0.05).
3. in a model including lTLE and rTLE patients (controlling for age, sex and dataset-site, epilepsy duration, existence of hippocampal sclerosis and spikes/minute), we observed a significant increase of FC_fMRI_-FC_EEG-δ_/-FC_EEG-θ_/-FC_EEG-α_/-FC_EEG-β_ correlation when comparing rTLE to lTLE patients (rTLE>lTLE, p<0.05/5=0.01, Bonferroni corrected)

For detailed results of the bootstrap analysis see SI Table 5.

### Local spatial contributions to the FC_fMRI_-FC_EEG_ correlation

We then compared the FC_fMRI_-FC_EEG_ correlation of subnetworks that take only into account connections from one specific ICN between TLE patients and healthy controls. We observed that when comparing lTLE patients to healthy controls the FC_fMRI_-FC_EEG-β_ correlation was significantly decreased for lTLE patients in the DMN (lTLE<controls, permutation of group labels, 5000 iterations, p<0.05/(5*7)=0.0014, corresponding to a Bonferroni threshold p<0.05, Fig 3B), while no significant alterations were observed when comparing rTLE patients to healthy controls (rTLE>controls, permutation of group labels, 5000 iterations, p>0.05/(5*7)=0.0014, corresponding to a Bonferroni threshold p>0.05).

When comparing the spatial contribution to the global EEG-fMRI connectome correlation (group averaged) between lTLE patients and healthy controls we observed a significantly decreased spatial contribution in FC_fMRI_-FC_EEG-β_ correlation inside the DMN for (see Fig 3B) (lTLE<controls, permutation of group labels, 5000 iterations, p<0.05/(5*7)=0.0014, corresponding to a Bonferroni threshold of p<0.05). When comparing rTLE patients with healthy controls we did not observe a shift in contribution to the global correlation for specific ICNs (rTLE>controls, permutation of group labels, 5000 iterations, p>0.05/(5*7)=0.0014, corresponding to a Bonferroni threshold of p>0.05).

## Discussion

This study based on simultaneously recorded EEG and fMRI functional connectivity in patients with temporal lobe epilepsy and healthy controls in two independent datasets, characterisedhow a whole-brain network approach in epilepsy relates between both modalities. We replicated the moderate relationship between whole-brain FC_fMRI_ and FC_EEG_ in healthy controls (Wirsich et al., 2021) and we confirmed for the first time that this relationship also exists in patients with epilepsy. While networks of rTLE patients show a widespread change of the relationship for the lower EEG frequency bands, the networks of lTLE patients have a global relationship of EEG and fMRI connectivity more similar to controls. Nevertheless, alterations between lTLE patients and healthy controls were observed locally (in particular the DMN) and were linked to FC_EEG-β_. This suggests that functional network reorganization across multiple timescales undergoes a more widespread or heterogeneous change in rTLE patients, impacting the relationship between EEG and fMRI, while alterations of the multimodal relationship are more homogenously localized in lTLE patients.

### Monomodal relationship

For both datasets and in line with our previous research (Wirsich et al., 2021) we showed that monomodal intragroup correlation was high (FC_fMRI_) to moderate (FC_EEG-γ_). When comparing connection-wise differences in networks between patients and controls we were unable to observe any significant differences between controls vs. lTLE patients and controls vs. rTLE patients. This is opposed to our previous findings in rTLE (Wirsich et al., 2016), where we observed FC_fMRI_ differences in rTLE patients vs. controls (though using a high resolution 512 regions atlas as opposed to the low-resolution atlas of 68 regions used in this study). The difficulty of identifying a consistent localized network across patients reflects the general heterogeneity of network neuroscience literature in epilepsy which can be very sensitive to individual methodological choices analyzing resting-state connectivity (Centeno and Carmichael, 2014; Slinger et al., 2022; van Diessen et al., 2014). In summary, we observed that spatial localization of monomodal FC differences lack a spatial homogeneity that can be detected with the small group-size of 34 patients used here. The heterogeneity of connection-wise alterations of individual connections observed here can potentially be mediated by describing the network topologically using graph theoretic descriptions (Carboni et al., 2020; Ridley et al., 2015; Wirsich et al., 2021, 2016).

### FC_fMRI_-FC_EEG_ correlation

The simplest way to compare brain networks derived from different modalities on a topological level is using the spatial correlation of the connectivity (Honey et al., 2009; Wirsich et al., 2017). We observed significantly increased global FC_fMRI_-FC_EEG_ correlation in rTLE as compared to healthy controls. Conversely global alterations between lTLE patients and controls were restricted in timescale to the FC_fMRI_-FC_EEG-β_ relationship which was found to be locally dominant in the Default Mode Network. This is in line with the observation of Coito et al. (2015) and Zhao et al. (2022) showing that FC_EEG_ of rTLE patients undergoes more widespread alterations in brain networks than those of lTLE patients.

This finding of functional alterations affecting regions remote to the epileptic focus resulting in global shift of functional networks altered by epilepsy as a function of the laterality of the epilepsy is further supported by a recent multicentric study showing that while atrophy in lTLE is more restrained to the ipsilateral side while in rTLE both ipsi and contralateral side are affected (Park et al., 2021). From a structural point of view, we previously observed that FC_fMRI_ of rTLE patients is more closely related to structural connectivity derived from diffusion MRI than healthy controls (Wirsich et al., 2016). Together with the results in the current study in rTLE patients, this points to a general increase of correlation between both structural and functional connectivity across all temporal scales. Interestingly for the FC_EEG_-FC_fMRI_ correlation this does not seem to be the case in lTLE patients. Future work should validate if this is also true for the structure-function relationship.

We previously demonstrated that FC_fMRI_ and FC_EEG_ hold both distinct and mutual information (Wirsich et al., 2020a, 2017). Though we observed a moderate correlation between FC_fMRI_ and FC_EEG_ for both healthy controls and TLE patients, this work confirms that FC_EEG_ and FC_fMRI_ studies do not measure exactly the same properties in line with the disconnect between FC_fMRI_ and FC_EEG_ graph analysis literature (Slinger et al., 2022). The results of our study stress that the relationship between FC_fMRI_ and FC_EEG_ is only partial and, more importantly, alters with the lateralization of epilepsy, limiting the direct comparability of EEG and fMRI connectome studies.

### Spatial contribution of FC_fMRI_-FC_EEG_ correlation

From a structural parcelation point of view, asymmetries between left and right temporal lobe have been widely described (Van Essen et al., 2012). Rather than a limitation of the functional repertoire (Wirsich et al., 2016) the differential spatial contributions in rTLE and lTLE patients suggest different adaptations of normal healthy functional networks to epilepsy e.g. more healthy bilateral functional integration of right temporal lobe vs. a more localized function of the left temporal lobe (Raemaekers et al., 2018). Looking exclusively at the DMN, Haneef et al. (2012) observed that local changes of fMRI connectivity are larger in lTLE as compared to rTLE. In line we observed that the decrease in EEG-fMRI connectivity relationship was linked locally to the DMN in lTLE but not in rTLE. We extend the observation of Haneef et al. (2012) by also showing that the multimodal connectivity reorganization is linked to a local change of FC_fMRI_-FC_EEG-β_ correlation in lTLE.

The cognitive consequences of differential reorganization in rTLE vs. lTLE are for example illustrated by the results of Drane et al. (2013), demonstrating that while rTLE patients have problems with recognizing famous faces, lTLE patients rather have problems naming them. From a physiological point of view the results are also in line with the general asymmetry of connectivity in temporal regions resulting in increased local connectivity in the left hemisphere when compared to the right hemisphere (Raemaekers et al., 2018).

### Implications for clinical research

While we looked only at temporal lobe epilepsy the observed lateralized discrepancy in the relationship of FC_EEG_ and FC_fMRI_ might not be limited to TLE but could apply more generally to the lateralization of the epileptic zone in epilepsy (Ridley et al., 2015). Further, this feature might not only be a sensitive marker restricted to epilepsy but it might be also linked to lateralization of brain dysfunction (e.g. one could see the same effect in lateralized tumors or strokes that alter the brain network). Further studies would be needed to better understand the relationship in EEG and fMRI in other focal neuro-pathologies.

Apart from lateralization of the TLE - using a bootstrapping approach - we did not observe that clinical parameters contribute significantly to the alteration of the EEG-fMRI relationship, suggesting that those parameters do modulate FC_fMRI_ and FC_EEG_ in a similar way between differently lateralized epilepsies. Consequently, the relationship of FC_fMRI_ and FC_EEG_ might provide a potential additional clinical marker to determine lateralization (Douw et al., 2019; Sadaghiani and Wirsich, 2020). Our results are encouraging as they generalize across two datasets and future work should validate if the EEG-fMRI can be clinically used to determine if a patient has a lateralization of epilepsy in the left or right hemisphere.

### Methodological considerations

From a spatial point of view, reconstructing EEG brain activity from deep cortical regions (such as the hippocampus) is still a subject of discussion (Pizzo et al., 2019). As such our approach to symmetrically integrate FC_fMRI_ and FC_EEG_ was limited to temporal lobe without the hippocampus as defined by Desikan et al. and (2006) and Yeo et al. (2011) but including neighboring cortical regions such as the parahippocampal gyrus and temporal pole As improving SNR of FC_EEG_ from hippocampal regions is still ongoing research, future work might profit form integrating monomodal FC_fMRI_ asymmetrically in this framework.

Further, while separating rTLE patients between the two recording sites, we demonstrated that results exist individually for each site. We included only three lTLE patients for the 64Ch-3T dataset, nevertheless, using the proposed bootstrap approach, we did not observe any systematic effects of dataset-site when pooling all the subjects together.

In this study we selected only clear cases of lateral temporal lobe epilepsy sampled out of a database of ∼200 EEG-fMRI recordings for the 256Ch3T dataset and ∼60 recordings for the 64Ch3T dataset to assure relative homogeneity of the groups. The final group of 34 patients was the most homogenous group with reasonable sample size. Though, when comparing TLE patients to controls, we demonstrated global changes of the FC_fMRI_ and FC_EEG_ relationship, we were unable to extract a common network of reorganization based on pair-wise connections (both for EEG and fMRI). Better understanding of individual functional networks linked to epilepsy beyond the group-averaged approach taken here (Marek et al., 2022; Wirsich et al., 2021, 2016) will need a larger database with data pooling in an even more multicentric approach (Marek et al., 2022; Slinger et al., 2022).

A larger multicentric approach would equally help to characterize the effect of individual antiseizure medication treatment on FC (Wandschneider and Koepp, 2016; Xiao et al., 2019) which was not taken into account. This effect is potentially negligible in our data as both rTLE and lTLE patients will undergo the comparable treatment and previously measured drug-effects on the EEG-fMRI correlation were observed to be small (Forsyth et al., 2019). However, a systematic characterization of a medication effect (Wirsich et al., 2018) is still missing in the research field of characterizing functional networks in focal epilepsies.

## Conclusions

In this study we investigated the FC_fMRI_-FC_EEG_ correlation in healthy controls and in TLE patients. We observed that monomodal alterations between controls and TLE are hard to track. However, when looking at the spatial correlation between FC_fMRI_ and FC_EEG_ we were able to demonstrate global alterations between rTLE patients and healthy controls, while alterations between lTLE patients and controls were more local. This demonstrates the differential organization of mono-lateral focal epilepsy of the same type that needs to be considered when comparing EEG to fMRI connectivity. It also demonstrates that each modality provides distinct information, highlighting the benefit of multimodal assessment in epilepsy. This property of distinct topological patterns depending on the lateralization of the epilepsy could be taken into account when clinically defining the epileptic focus of patients.

## Data Availability

All anonymized connectomes are available on Zenodo (10.5281/zenodo.7025003). Raw data can be made fully available on request within the limits of ethical regulations of Switzerland and France. As such the project of the requesting party will need to undergo a formal ethical approval procedure (on request to SV and 256Ch-3T dataset or to MG for the 64Ch-3T dataset). Code is available at Github (https://github.com/jwirsich/eeg-fmri-tle).

doi://10.5281/zenodo.7025003

https://github.com/jwirsich/eeg-fmri-tle

## Acknowledgments

We acknowledge support from Swiss National Science Foundation (SNSF, under grants CRSII5_209470, and 192749 to SV, 163398 and 180365 to MS and 188769 to FG, P500PM_206720 to LS). JW is supported by a research position of the Faculty of Medicine, University of Geneva. This work was supported by Centre d’Imagerie BioMédicale (CIBM) of the UNIL, UNIGE, HUG, CHUV, EPFL and the Leenaards and Jeantet Foundations, the CNRS, the ANR CONNECTEPI (grant number ANR-07-NEUR-0010).

## Supplementary Information

**SI Table 1:** Scanner setup for both centers. The scanner of the 256Ch-3T dataset received a scanner update during acquisition (data from years 2010-2019), which resulted in a slight change of acquisition parameters

| Dataset | Scanner | TR | Resolution | Number of controls | Number of patients |
| --- | --- | --- | --- | --- | --- |
| **256Ch-3T** | Siemens Trio | 1980ms | 3x3x3.75mm | 0 | 8 |
| **256Ch-3T** | Siemens Trio | 1990ms | 3x3x3.75mm | 17 | 10 |
| **256Ch-3T** | Siemens Trio | 2000ms | 3x3x3.75mm | 0 | 1 |
| **256Ch-3T** | Siemens Prisma | 1990ms | 3x3x3.75mm | 4 | 4 |
| **64Ch-3T** | Siemens Magnetom Verio | 3600ms | 2.0x2.0x2.5mm | 14 | 11 |

**SI Table 2:** Splithalf and crossdataset spatial correlation of monomodal FC_EEG_ and FC_fMRI_ (random permutations of splithalf assignments with 5000 iterations, in case of n<16 all possible combinations to split the group into two halves).

|  | fMRI | delta | theta | alpha | beta | gamma |
| --- | --- | --- | --- | --- | --- | --- |
| **Controls** |  |  |  |  |  |  |
| Split half 256Ch3T | 0.87054 | 0.73682 | 0.77674 | 0.75082 | 0.82411 | 0.49047 |
| Split half 64Ch3T | 0.87817 | 0.80862 | 0.7131 | 0.56457 | 0.57558 | 0.35359 |
| Cross dataset | 0.77132 | 0.73061 | 0.66702 | 0.64033 | 0.68208 | 0.4403 |
| **ITLE** |  |  |  |  |  |  |
| Split half 256Ch3T | 0.78767 | 0.71863 | 0.7437 | 0.75605 | 0.82403 | 0.58585 |
| **rTLE** |  |  |  |  |  |  |
| Split half 256Ch3T | 0.62129 | 0.679 | 0.7551 | 0.75985 | 0.80257 | 0.56043 |
| Split half 64Ch3T | 0.78417 | 0.58941 | 0.46924 | 0.37836 | 0.48847 | 0.28072 |
| Cross dataset | 0.66777 | 0.65887 | 0.58191 | 0.45151 | 0.59768 | 0.25093 |

**SI Table 3:**
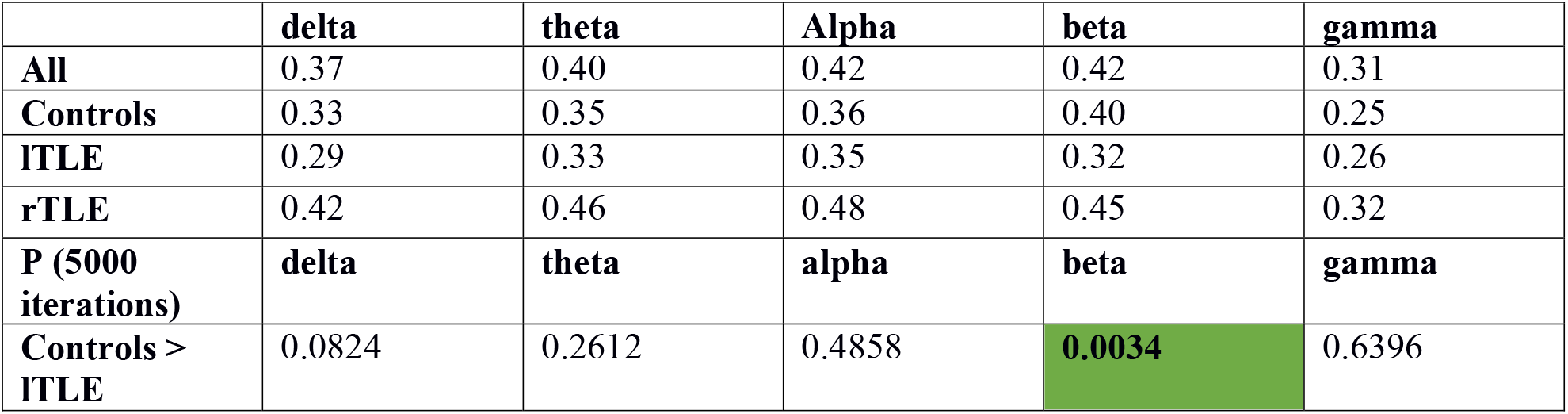
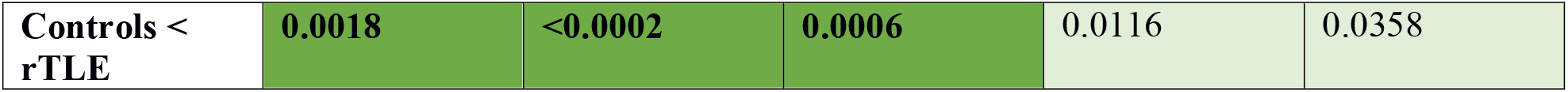
Spatial correlation between FC_fMRI_ and FC_EEG_ for each frequency band averaged for each group across both datasets (5000 permutations of group labels, significant results marked in dark green bold: Bonferroni threshold p<0.05/5=0.01), Light green: Uncorrected threshold p<0.05.

|  | delta | theta | Alpha | beta | gamma |
| --- | --- | --- | --- | --- | --- |
| **All** | 0.37 | 0.40 | 0.42 | 0.42 | 0.31 |
| **Controls** | 0.33 | 0.35 | 0.36 | 0.40 | 0.25 |
| **ITLE** | 0.29 | 0.33 | 0.35 | 0.32 | 0.26 |
| **rTLE** | 0.42 | 0.46 | 0.48 | 0.45 | 0.32 |
| **P (5000 iterations)** |  |  |  |  |  |
| **Controls > ITLE** | 0.0824 | 0.2612 | 0.4858 | **0.0034** | 0.6396 |
| **Controls < rTLE** | **0.0018** | **<0.0002** | **0.0006** | 0.0116 | 0.0358 |

**SI Table 4:** Spatial correlation between FC_fMRI_ and FC_EEG_ for each frequency band for each dataset (rows for lTLE/64Ch-3T are not reported as only group is consisting of 3 patients, 5000 permutations of group labels, significant results marked in dark green bold: Bonferroni threshold p<0.05/5=0.01), Light green: Uncorrected threshold p<0.05.

|  | delta | theta | Alpha | beta | gamma |
| --- | --- | --- | --- | --- | --- |
| **256Ch-3T** |  |  |  |  |  |
| Controls | 0.31 | 0.33 | 0.35 | 0.37 | 0.23 |
| ITLE | 0.28 | 0.32 | 0.33 | 0.31 | 0.23 |
| rTLE | 0.38 | 0.39 | 0.41 | 0.39 | 0.31 |
| **64Ch-3T** |  |  |  |  |  |
| Controls | 0.29 | 0.27 | 0.22 | 0.29 | 0.14 |
| rTLE | 0.36 | 0.38 | 0.31 | 0.33 | 0.15 |
| **P (5000 iterations)** | **delta** | **theta** | **alpha** | **beta** | **gamma** |
| **256Ch-3T** |  |  |  |  |  |
| Controls > ITLE | 0.4024 | 0.561 | 0.5904 | 0.066 | 0.6464 |
| Controls < rTLE | **0.0046** | 0.0116 | 0.0124 | 0.077 | 0.0114 |
| **64Ch-3T** |  |  |  |  |  |
| Controls < rTLE | **<0.002** | **<0.002** | **<0.002** | **<0.002** | 0.0876 |

**SI Table 5:** P-Values of permutation test with 5000 iterations of linear model coefficients when bootstrapping averages on contrast of interest controls vs. lTLE/rTLE (dark green Bold: Bonferroni threshold p<0.05/5=0.01, Light green: Uncorrected threshold p<0.05, contrast of interest marked in light gray) and lTLE vs. rTLE (dark green Bold: Bonferroni threshold p<0.05/5=0.01, Light green: Uncorrected threshold p<0.05, only for contrast of interest marked in light gray). The Permutation test is carried out by switching labels of interest. Significant values are marked in BOLD italics. HS: hippocampal sclerosis

|  | delta | theta | alpha | beta | gamma |
| --- | --- | --- | --- | --- | --- |
| **Controls & ITLE** |  |  |  |  |  |
| Age | 0.4400 | 0.1514 | 0.1478 | 0.3968 | 0.2268 |
| Sex | 0.0022 | 0.0220 | 0.0284 | 0.0762 | 0.0938 |
| Controls vs. ITLE | 0.4970 | 0.3700 | 0.2966 | 0.0450 | 0.1696 |
| 256Ch-3T vs. 64Ch-3T | 0.2598 | 0.4700 | 0.0242 | 0.4244 | 0.2220 |
| **Controls & rTLE** |  |  |  |  |  |
| Age | 0.1542 | 0.2026 | 0.2802 | 0.1226 | 0.2202 |
| Sex | 0.0060 | 0.0246 | 0.1042 | 0.0530 | 0.1130 |
| Controls vs. rTLE | **0.0002** | **<0.0002** | **<0.0002** | **0.0016** | 0.0404 |
| 256Ch-3T vs. | 0.2930 | 0.3222 | 0.0782 | 0.2800 | 0.0062 |
| 64Ch-3T |  |  |  |  |  |
| **ITLE & rTLE** |  |  |  |  |  |
| Age | 0.3616 | 0.0728 | 0.0298 | 0.4282 | 0.3222 |
| Sex | 0.4834 | 0.2930 | 0.1266 | 0.4722 | 0.0534 |
| HS vs. Non-HS | 0.2734 | 0.0118 | 0.0080 | 0.0064 | 0.0176 |
| Epilepsy Duration | 0.2304 | 0.3490 | 0.2738 | 0.3706 | 0.1794 |
| AED/Min | 0.2268 | 0.1000 | 0.3008 | 0.0936 | 0.1282 |
| ITLE vs. rTLE | **0.0002** | **0.0014** | **0.0052** | **0.0002** | 0.0272 |
| 256Ch-3T vs. 64Ch-3T | 0.3730 | 0.2926 | 0.1236 | 0.4280 | 0.0244 |

**SI Table 6:**
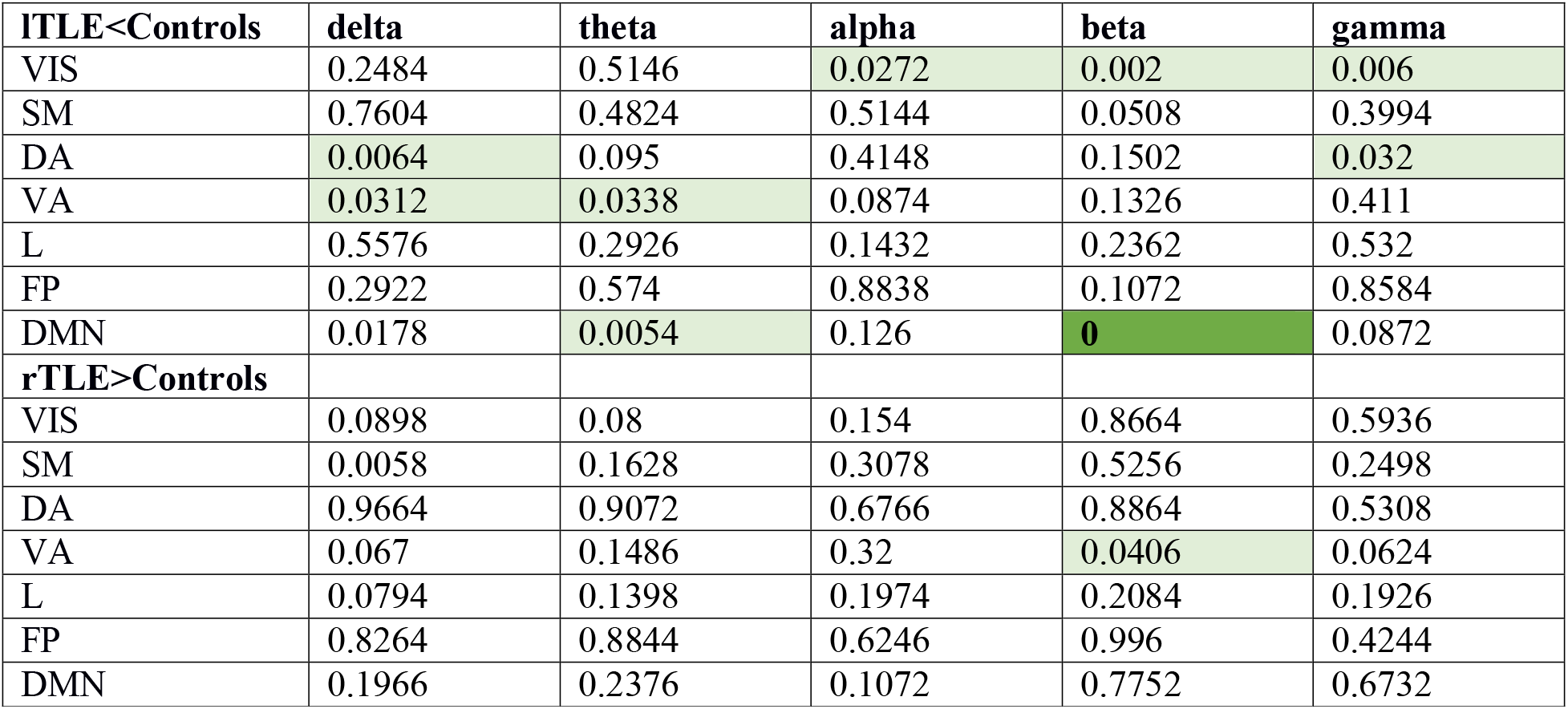
Comparison between TLE patients and healthy controls for EEG-fMRI correlation restricted to the connections of inside each intrinsic connectivity networks (Yeo et al., 2011). Dark green: Bonferroni threshold p<0.05/(7*5)∼0.0014, Light green: Uncorrected threshold p<0.05.

**SI Table 7:** Comparison between TLE patients and healthy controls for the spatial contribution (Colclough et al., 2016) of intrinsic connectivity networks (Yeo et al., 2011) to the global EEG-fMRI correlation. Dark green: Bonferroni threshold p<0.05/(7*5)∼0.0014, Light green: Uncorrected threshold p<0.05.

| ITLE<Controls | delta | theta | alpha | beta | gamma |
| --- | --- | --- | --- | --- | --- |
| VIS | 0.2292 | 0.3922 | 0.0288 | 0.1688 | 0.1402 |
| SM | 0.3844 | 0.7714 | 0.9978 | 0.2588 | 0.1412 |
| DA | 0.3224 | 0.5842 | 0.7258 | 0.2026 | 0.0968 |
| VA | 0.5304 | 0.6254 | 0.9978 | 0.5534 | 0.0506 |
| L | 0.9814 | 0.8436 | 0.6298 | 0.9852 | 1 |
| FP | 0.9872 | 0.995 | 0.9302 | 0.7752 | 0.8668 |
| DMN | 0.0062 | 0.0052 | 0.007 | 0.001 | 0.2062 |
| rTLE>Controls |  |  |  |  |  |
| VIS | 0.822 | 0.6796 | 0.8986 | 0.9562 | 0.7908 |
| SM | 0.0846 | 0.0266 | 0.0314 | 0.4296 | 0.8552 |
| DA | 0.6682 | 0.54 | 0.6578 | 0.931 | 0.4088 |
| VA | 0.0188 | 0.0096 | 0.0224 | 0.1516 | 0.849 |
| L | 0.37 | 0.7082 | 0.7974 | 0.1312 | 0.0088 |
| FP | 0.2856 | 0.0618 | 0.2184 | 0.7044 | 0.5238 |
| DMN | 0.875 | 0.9488 | 0.8214 | 0.654 | 0.9562 |

